# Harmonization of Real-World Data for Vaccine-Preventable Infectious Diseases: Integration of SARS-CoV-2 Diagnostic and Serologic Data from Multiple Sources

**DOI:** 10.64898/2026.01.14.26344023

**Authors:** Yonah C. Ziemba, Suhyeon Yoon, Harvey W. Kaufman, William A. Meyer, Laura Gillim, Nkemakonam Okoye, Cheryl B. Schleicher, Ligia A. Pinto, Lynne Penberthy, James M Crawford

**Author notes:** Corresponding Author: James M. Crawford, M.D., Ph.D. Department of Pathology and Laboratory Medicine 1111 Marcus Avenue, Suite 104B New Hyde Park, NY 11042.

## Abstract

**Background:** The COVID-19 Real World Data infrastructure (CRWDi) was established to study the impact of COVID-19 on patients with immunocompromising conditions. A key challenge was harmonizing SARS-CoV-2 molecular and serologic laboratory test results. *Methods.* There were 27 different test names for SARS-CoV-2 nucleic acid amplification tests (NAAT); and 34 and 26 for SARS-CoV-2 qualitative and semi-quantitative serologic tests, respectively. We validated a recoding strategy to eliminate the multiplicity of test names and results, and harmonized SARS-CoV-2 semi-quantitative serology test results by target antigen and antibody; published conversion factors were used to report data on a uniform Binding Arbitrary Units (BAU) scale.

**Results:** For 5 200 000 patients, the number of unique SARS-CoV-2 test events were: 4 865 431 NAAT; 3 092 198 qualitative; and 834 487 semi-quantitative serology tests, of which 378 522 were anti-spike (S) semi-quantitative serology. In achieving harmonization of semi-quantitative serologic results, semilog cumulative ordinal plots demonstrated that test results from patients with transplantation exhibited a higher proportion of negative-and-submaximal SARS-CoV-2 anti-S BAU values, when compared to patients with cancer or systemic autoimmune or rheumatic diseases (SARDs), or to a general non-immunocompromised population.

**Conclusions:** Complex and heterogeneous test name and result conventions represent a significant barrier to understanding real-world laboratory data for SARS-CoV-2. The generalizable methodology described herein permits successful harmonization of test data from multiple laboratory sources with administrative claims data, vital status data, and structured data on cancer diagnostics and treatment. This methodology supports exploration of the utility of semi-quantitative serologic data in all patients, especially those with immunocompromising conditions.

**Summary:** We present a strategy for harmonizing real-world laboratory test results, based on the COVID-19 Real World Data infrastructure (CRWDi). By successfully addressing this barrier, this strategy is of general value for studying the clinical utility of laboratory testing.

## Introduction

Infectious diseases are part of our species’ existence. Regardless of whether pathogens are previously identified or are novel, tracking their impact on human populations is essential for management of public health and for informing medical practice (1). Well-established national and global public surveillance networks can adapt quickly to the detection and monitoring of emergent novel pathogens (2). However, the recent COVID-19 pandemic demonstrated the challenges of having an extant data infrastructure to develop real-world evidence to inform medical practice. Fortunately, productive multi-institutional consortia were quickly established that addressed key data needs for understanding and managing the COVID-19 pandemic (3,4), including tracking the efficacy of vaccination for administration to the public (5,6).

However, important questions persisted through the pandemic including the impact of COVID-19 on immunocompromised patients (7,8), and the utility of SARS-CoV-2 serologic testing in assessing the host immune response following either natural infection or vaccination (9,10). In the first instance, United States (US)-based studies have provided strong evidence regarding the risk of immunocompromised patients for severe illness and post-acute sequelae of COVID-19 illness (11–14), and the importance of vaccination (including booster vaccination) for reducing the risk of severe COVID-19 illness (15–17). In the second instance, however, SARS-CoV-2 anti-spike protein (anti-S) serologic testing was not endorsed in the US as having diagnostic or prognostic utility other than in assessing children and adults with potential multisystem inflammatory syndrome (18), despite detailed data on host immune response in immunocompetent and immunocompromised populations (19). This perceived lack of utility for SARS-CoV-2 anti-S serologic testing is in contrast to the long-documented value of serologic testing for surveillance of other vaccine-preventable pathogens such as measles virus, mumps, hepatitis B virus, and varicella, (20), for evaluation of patient immune status (21,22), and for guiding care of immunocompromised patients as regards their need for primary or booster vaccination (23–25).

The COVID-19 Real World Data infrastructure (CRWDi) was funded by the National Cancer Institute (NCI) to address gaps in knowledge about the impact of COVID-19 on immunocompromised patients, either based on their underlying condition and/or because of immune-modulating therapies (26). Established in May 2024 and containing 5.2 million unique patient records, the CRWDi data infrastructure is a de-identified, fully harmonized linkage of medical and pharmacy administrative claims data, vital status data, COVID-19 vaccination status data, and SARS-CoV-2 diagnostic and serologic laboratory data, with linkage also to deidentified patient records from the NCI Surveillance, Epidemiology, and End Results (SEER) network. Following an update in 2025, CRWDi spans the COVID-19 pandemic for calendar years 2020 to 2024. Owing to the combined contribution of national data from the two largest national commercial laboratories, another commercial laboratory, and regional data from the largest health system in the greater New York metropolitan area, CRWDi constitutes a robust repository of SARS-CoV-2 laboratory testing data that, in particular, provides a unique opportunity to address the utility of serologic testing in the assessment of patients made vulnerable through cancer, systemic autoimmune and rheumatic diseases (SARDs), and solid organ or stem cell transplantation. We report our strategies for successfully harmonizing these real-world laboratory data, with the goal of these strategies having general applicability for real-world study of the medical and public health utility of serologic testing.

## Methods

### Source SARS-CoV-2 laboratory test data

Two major commercial laboratories (Labcorp, Burlington, NC; Quest Diagnostics, Secaucus, NJ) contributed SARS-CoV-2 testing data, along with an integrated delivery network (Northwell Health, New Hyde Park, NY), and a third commercial source for SARS-CoV-2 NAAT testing only that was not disclosed to the authors. The inclusion dates for SARS-CoV-2 test data were March 1, 2020 to December 31, 2024. SARS-CoV-2 diagnostic testing was qualitative nucleic acid amplification testing (NAAT). Cycle threshold (Ct) values were not reported, and no antigen-based testing was included. SARS-CoV-2 serologic testing reported from the 3 identified laboratories included qualitative results for all test specimens and semi-quantitative serologic test values on a subset of specimens.

### Normalization and harmonization of SARS-CoV-2 laboratory data

The CRWDi data infrastructure is organized into relational tables; the laboratory relational tables include data fields for claim identification number, HealthVerity identification number (HVID), diagnostic codes entered on laboratory test requisitions, performing laboratory, date of test performance, test ordered name, test Logical Observation Identifiers Names and Codes (LOINC, Regenstrief Institute, Indianapolis, IN), result name, result unit of measure, reference range, result, and result comment.

#### Normalization of Test Name

The fundamental test categories were SARS-CoV-2 NAAT and SARS-CoV-2 serologic testing for total antigen, anti-spike (S) or anti-nucleocapsid (N) antigens. Antibody class was either total antibody, IgG, IgM, or IgA. Although LOINC codes provided an initial scheme for identifying test type, substantial variation in test names remained. At the beginning of the pandemic, the SARS-CoV-2 serologic test was named “SARS-CoV-2.” However, as assays were further developed, tests were named with more specific designation of whether the detected antibody was “anti-S” or “anti-N.” In similar fashion, in the early months of the pandemic the serologic assays detected total immunoglobulin; specification of antibody class (IgG, IgM, and IgA) became available as different manufacturers stood up antibody class-specific assays and received regulatory approval. **Supplemental Table S1** shows the multiplicity of test names requiring normalization (NAAT, n=27; qualitative serology, n=34; semi-quantitative serology, n=26).

#### Deduplication of test results

Following test name normalization, all test categories still had replicate entries for the same testing events. For any given test result, the International Disease Classification Version 10 (ICD-10) codes entered by the ordering physician on the test requisition were retained as unique data elements but populated separate rows for each test event. Hence, a test requisition with only one accompanying ICD-10 code was not replicated; a test requisition with 2 accompanying ICD-10 codes was reported in duplicate, and so on. **Table 1** shows the breakdown of replicated SARS-CoV-2 test results in CRWDi, for each test category. This coding logic is available to researchers using CRWDi to ensure that such replication is removed from future analytic work.

**Table 1.** Deduplication of SARS-CoV-2 test results. Data is presented for all test categories from the 4 source laboratories. The seven cardinal data fields that described a unique test result were: HealthVerity deidentified patient record number (HVID); administrative claim number; source laboratory; date of test; test name; test units; and test reference range (including for qualitative testing). However, replication of test results in CRWDi occurred due to multiple ICD-10 codes entered at the time of test ordering; each ICD-10 code generated a separate entry (row) for the test with which it was associated. Deduplication of test events (and hence rows in the relational table) required: (a) identifying unique test events using the seven cardinal data fields; (b) deduplicating multiple rows associated with single test events but which were created for each ICD-10 code on the laboratory test requisition; (c) generating a deduplicated derivative table for SARS-CoV-2 laboratory test results in the CRWDi data infrastructure. (The source data table was retained, so that these administrative claims data were not lost and remain available for analysis.) This table shows the number of unique tests with singlicate rows in the source laboratory test table, those with duplicate entries, etc. Deduplication of this magnitude underscores the degree of systematic pre-processing of real-world data necessary to avoid inflating counts.

| Rows Per Result | Total rows | NAAT distinct results | Serology qualitative distinct results | Serology semi-quantitative distinct results | Total distinct results |
| --- | --- | --- | --- | --- | --- |
| 1 | 6,497,860 | 3,581,269 | 2,260,204 | 656,387 | 6,497,860 |
| 2 | 3,143,262 | 1,127,334 | 397,696 | 46,601 | 1,571,631 |
| 3 | 626,595 | 75,601 | 113,939 | 19,325 | 208,865 |
| 4 | 564,620 | 53,523 | 71,613 | 16,019 | 141,155 |
| 5 | 436,360 | 11,983 | 59,054 | 16,235 | 87,272 |
| 6 | 395,694 | 5,810 | 45,074 | 15,065 | 65,949 |
| 7 | 360,276 | 2,544 | 35,507 | 13,417 | 51,468 |
| 8 | 323,544 | 2,063 | 27,309 | 11,071 | 40,443 |
| 9 | 289,611 | 1,163 | 21,449 | 9,567 | 32,179 |
| ≥10 | 1,137,521 | 4,141 | 60,353 | 30,800 | 95,294 |
| **Total** | **13,775,343** | **4,865,431** | **3,092,198** | **834,487** | **8,792,116** |

#### Parsing of test categories

We next established logic for test results categorization (**Table 2**): qualitative assays were first identified (SARS-CoV-2 NAAT and SARS-CoV-2 qualitative serologic testing), and thence SARS-CoV-2 semiquantitative serologic testing. The number of unique, deduplicated test results for each unique test category are given.

**Table 2.**
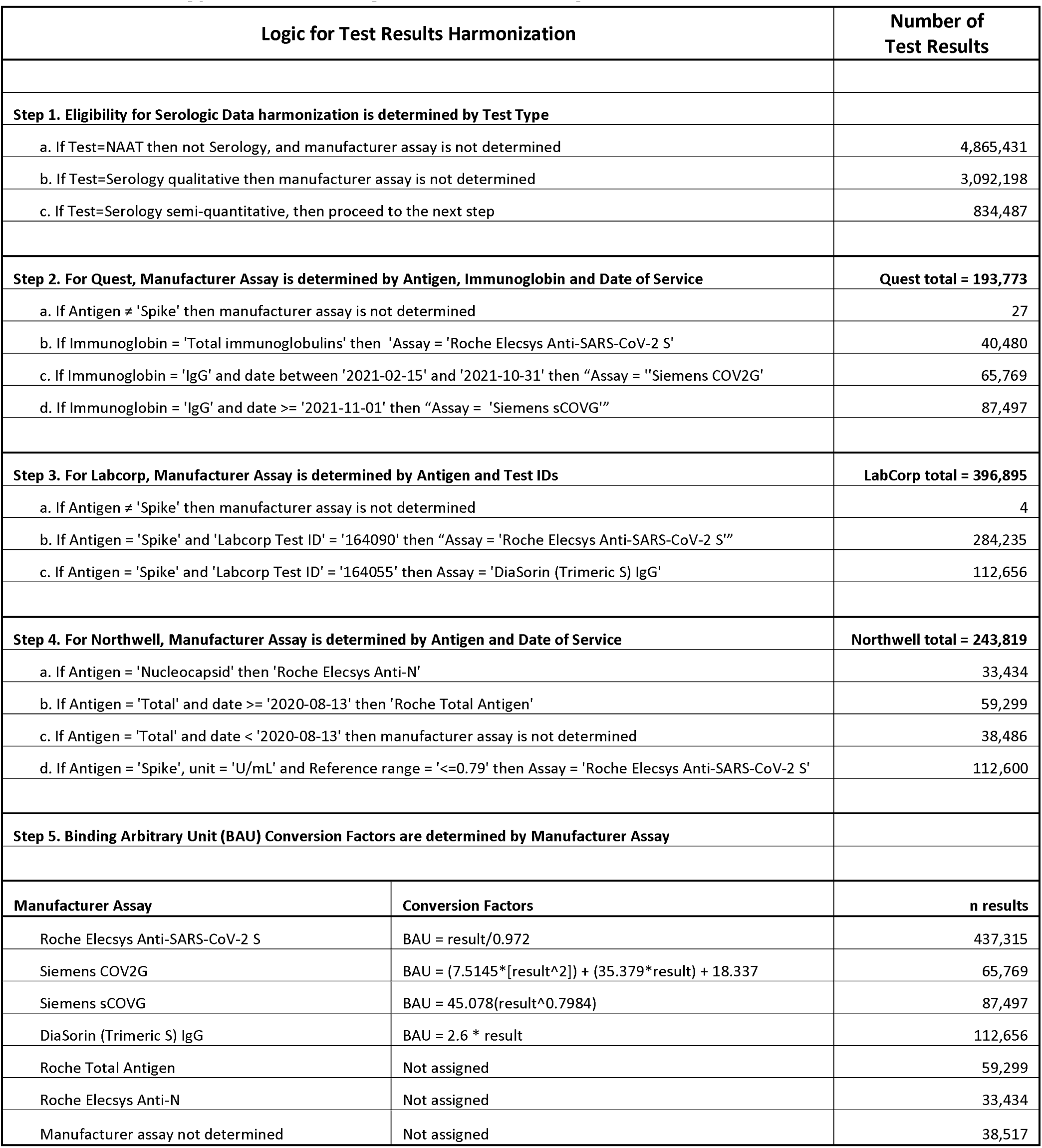
Strategy for harmonizing SARS-CoV-2 testing data. Data is given for all test categories from the source laboratories. The first step was to identify test category; if NAAT or qualitative SARS-CoV-2 serology (steps 1a, 1b, respectively), then the test manufacturer did not need to be known. If semi-quantitative SARS-CoV-2 serologic test (step 1c), then use laboratory source to identify assay manufacturer. For Quest and Northwell, the calendar date of test performance was sufficient. For Labcorp, a decode table was required (see step 3). When the logic of step 1c yielded “null” for manufacturer test platform, harmonization of semi-quantitative numerical results could not be performed. When the test category was a SARS-CoV-2 anti-S assay, and the test result was associated with a known manufacturer test platform, the published conversion factors for SARS-CoV-2 Anti-S testing in step 4 enabled harmonization of numerical values (Kaufman HW 2023).

#### Normalization of qualitative test results

Having parsed the test categories, there was substantive variability in how qualitative tests were reported. For both NAAT or serologic testing, the qualitative result should ostensibly be “negative” or “positive.” However, the alphanumeric characters in the results field were variable, with 59 different ways to report one of these two results (**Supplemental Table S2**). A complete inventory of all potential alphanumeric text sequences in the results field were compiled and verified with the source laboratories for their corresponding assignment to “negative” or “positive”; all qualitative results were recoded to the appropriate outcomes.

#### Harmonization of SARS-CoV-2 serology test results

**Table 2** then outlines our strategy for harmonizing semi-quantitative serology test results: (1) identify target antigen (“total”, “spike”, “RBD”, or “nucleocapsid”); (2) identify antibody class (“total”, “IgG”, “IgM”, or “IgA”); (3) identify serologic test type (“qualitative” or “semiquantitative”); (4) if semi-quantitative, identify unit of measure (“null”, “U/ml”, or Binding Arbitrary Unit; “BAU”). For semi-quantitative serologic testing; we then: (5) identified the manufacturer’s reference range (semi-quantitative numerical threshold) for “negative” versus “positive” qualitative resulting; and (6) based on CRWDi data, identified the dynamic range of the assay (maximum reported numerical values) from each source laboratory. Semi-quantitative test results above the reported dynamic range as defined by a particular manufacturer were reported as “greater than.” To apply conversion factors for harmonizing the numerical test results of semi-quantitative testing, explicit assay identity was needed. For the two national commercial laboratories, this information was available in the results comment data field. This field was searched for manufacturer and assay names used during the study period. The natural language processing Arabic character sequences that were searched were: “diasori” for Diasorin Inc. (Stillwater, MN); “roche” for Roche Diagnostics (Indianapolis, IN), and “siemens” for Siemens Healthineers (Erlangen, Germany). The serologic assays for these manufacturers are given in **Supplemental Table S3**. The CRWDi relational table for commercial laboratory semi-quantitative test results was recoded to include a structured data field for each respective manufacturer, for each test result. The test name, manufacturer, and date of service were then used to identify the reference range in use at that time, and published conversion factors were used to obtain harmonized BAU (27).

For testing performed by Northwell Health, known after August 21, 2020, all SARS-CoV-2 serologic testing used the Roche platform. Unfortunately, although semi-quantitative serologic testing had been performed starting April 14, 2020, it was on multiple manufacturer platforms until early August 2020 (28), information which was not included in the CRWDi data infrastructure. Northwell Health data reported herein is thus only for results obtained using the Roche platform, August 21, 2020 to December 31, 2024.

### Data analysis

Using SQL programming language in the Databricks analytic environment of CRWDi (Databricks Inc., San Francisco, CA), harmonized SARS-CoV-2 test data was joined with recode reference tables to specify the following: immunocompromised status (non-immunocompromised; patients with cancer as identified by the SEER data registry; patients with SARDs as identified by ICD-10 coding who had received disease-relevant pharmaceutical therapy within the past year; and patients who were not already included in the SEER-identified cancer cohort but who had undergone solid organ or stem cell transplantation based on ICD-10 codes; 26); test category; assay manufacturer; qualitative result (negative or positive); and calendar year and month the test was performed.

For semi-quantitative SARS-CoV-2 serologic test data, additional recode fields included: immunoglobulin class (total, IgG, IgM, IgA) and target antigen (S, RBD, N protein); S and RBD target antigens were grouped as S antigen. Results were visualized using R software (The R Project for Statistical Computing, R Foundation, PO Box 776, Kaysville, UT 84037-0776) and Excel (Microsoft, Redmond, WA).

### Data Availability for Researchers

The CRWDi data resource was openly available before initiation of the study. The CRWDi data resource was developed to support academic, non-commercial research projects in the US (26). Information about this data resource can be found at (https://seer.cancer.gov/data-software/crwdi/). Per this website, this initiative is no longer accepting new data requests. The data infrastructure is to remain active for current users through its planned duration (April 2026). The aggregate datasets analyzed during the current study are available from the corresponding author on reasonable request.

### Patient Consent Statement

The Institutional Review Board of Northwell Health approved the performance of this study, under an exempt protocol and without requirement for patient consent. For proposed studies using CRWDi, each research investigator must obtain and show proof of institutional review board approval before the NCI will authorize HealthVerity to grant access to CRWDi.

## Results

### SARS-CoV-2 test volumes

There are a total of 8 792 116 unique SARS-CoV-2 testing events in the CRWDi data resource (**Table 3**), shown by month and year in **Figure 1A**. The preponderance of testing events was in calendar years 2020-2022. For SARS-CoV-2 serologic testing, qualitative predominated in 2020 to mid-2021. From November 2021 onwards, semi-quantitative testing usually accompanied qualitative test reporting.

**Figure 1.**
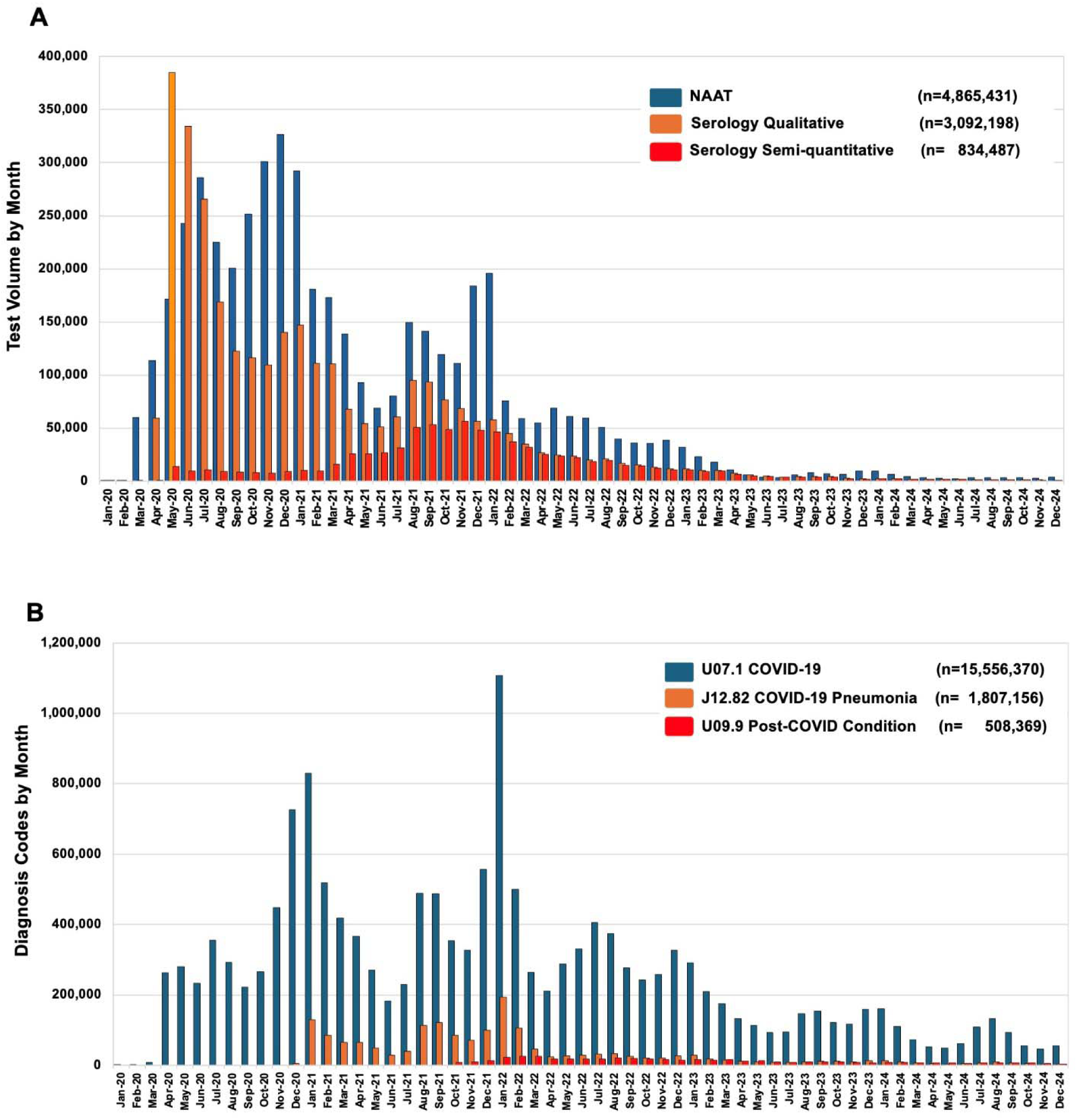
SARS-CoV-2 test volumes and COVID-19 ICD-10 coding volumes, 2020 to 2024. (A) Test volumes by month and calendar year are shown for nucleic acid amplification testing (NAAT), qualitative serology, and semi-quantitative serology; the legend gives total unique events for each test category. (B) Unique coding events by the International Classification of Diseases Tenth Revision (ICD-10) by month and calendar year are shown for the 5.2 million unique deidentified patients in the COVID-19 Real World Data infrastructure (CRWDi); the legend gives total unique coding events by ICD-10 code.

**Table 3.** SARS-CoV-2 test volumes by source laboratory, test category, and test subcategory. Following the deduplication shown in **Table 2**, the unique test events in the CRWDi data infrastructure are shown. Northwell, Quest, and LabCorp reported all three categories of test results; one other laboratory source reported only qualitative SARS-CoV-2 NAAT testing and was not included in the methodology for harmonizing SARS-CoV-2 serologic testing data.

| Test category | Northwell | Quest | Labcorp | Other | Total |
| --- | --- | --- | --- | --- | --- |
| **NAAT** | 416,975 | 1,717,297 | 1,786,820 | 944,339 | 4,865,431 |
| **Serology qualitative** | 243,777 | 1,039,579 | 1,808,842 |  | 3,092,198 |
| **Total IgG** | 97,538 | 840,460 |  |  | 937,998 |
| **Total IgM** |  | 54,359 |  |  | 54,359 |
| **Isotype not provided** |  | 303 |  |  | 303 |
| **Spike Total** | 112,833 |  | 260,024 |  | 372,857 |
| **Spike IgG** | 3 | 40 | 1,046,443 |  | 1,046,486 |
| **Spike IgM** |  | 55,353 | 287,770 |  | 343,123 |
| **Spike IgA** |  |  | 75,991 |  | 75,991 |
| **Nucleocapsid Total** | 33,403 | 397 | 138,614 |  | 172,414 |
| **Nucleocapsid IgG** |  | 88,667 |  |  | 88,667 |
| **Serology semi-quantitative** | 243,819 | 193,773 | 396,895 |  | 834,487 |
| **Total IgG** | 97,785 | 16 |  |  | 97,801 |
| **Spike Total** |  | 40,480 | 284,235 |  | 324,715 |
| **Spike IgG** | 112,600 | 153,266 | 112,656 |  | 378,522 |
| **Nucleocapsid Total** |  |  | 4 |  | 4 |
| **Nucleocapsid IgG** | 33,434 | 4 |  |  | 33,438 |
| **Total IgM** |  | 7 |  |  | 7 |
| **Total** | 904,571 | 2,950,649 | 3,992,557 | 944,339 | 8,792,116 |

### COVID-19 ICD-10 coding events

**Supplemental Table S4** shows ICD-10 codes relevant to COVID-19 illness that accompanied the SARS-CoV-2 test requisitions. The predominant ICD-10 codes were the following: asymptomatic screening (n=584 901) including screening for COVID-19 (Z11.52, n=47 386); contact (n=1 624 364) including exposure to COVID-19 (Z20.822, n=483 708); and symptoms or disease condition (n=500 454) including COVID-19 illness (U07.1, n=114 195). These potentially relevant ICD10 codes accompanying SARS-CoV-2 test orders (n=2 709 719) are far fewer in number than the total testing events (n=8 792 116).

**Figure 1B** shows the total ICD-10 coding in the CRWDi database, as obtained from administrative claims data and from laboratory test requisitions, by month and year. Three COVID-19 specific codes are shown: U07.1 (COVID-19 illness, n=15 556 370), J12.82 (COVID-19 pneumonia, n=1 807 156), and U09.9 (post-COVID condition, n=508 369). The vast preponderance of the 17 477 721 total ICD-10 codes for COVID-19 (U07.1, J12.82, U09.9) were thus from administrative claims data.

### Semi-quantitative SARS-CoV-2 Anti-S serologic testing

Harmonization of semi-quantitative SARS-CoV-2 anti-S serologic testing enabled visualization of semi-quantitative test results using cumulative frequency semi-logarithmic plots. The purpose of this visualization was to provide immediate appreciation of the percentage of test results that were negative, sub-maximal (falling within the dynamic range of the semi-quantitative assay), and maximal (above the dynamic range). Figure 2A shows the cumulative percentage of results from the 3 laboratories contributing serologic data. The dynamic range was 0-250 BAU for Northwell Health test results, for testing performed entirely on the Roche platform and without autodilution of specimens that resulted >250 BAU. This chart documents that Quest routinely performed 1:10 autodilution, with a harmonized dynamic range of 0-2500 BAU; the slightly variable plateaus represent different conversion factors for the different manufacturer platforms. In turn, Labcorp performed both 1:10 and 1:100 autodilutions, as documented by the plateaus both in the 2500 and 25 000 BAU range. Despite these differing autodilution practices between the 3 source laboratories, the data show that the percent of test results that were negative were comparable for the 3 laboratories. For Quest and Labcorp, anti-S antibody curves were similar up to the 1:10 dilution plateaus of approximately 2500 BAU. Lastly, for those Labcorp test specimens that were diluted 1:100, only a limited percentage resulted at the maximum of 25 000 BAU.

**Figure 2.**
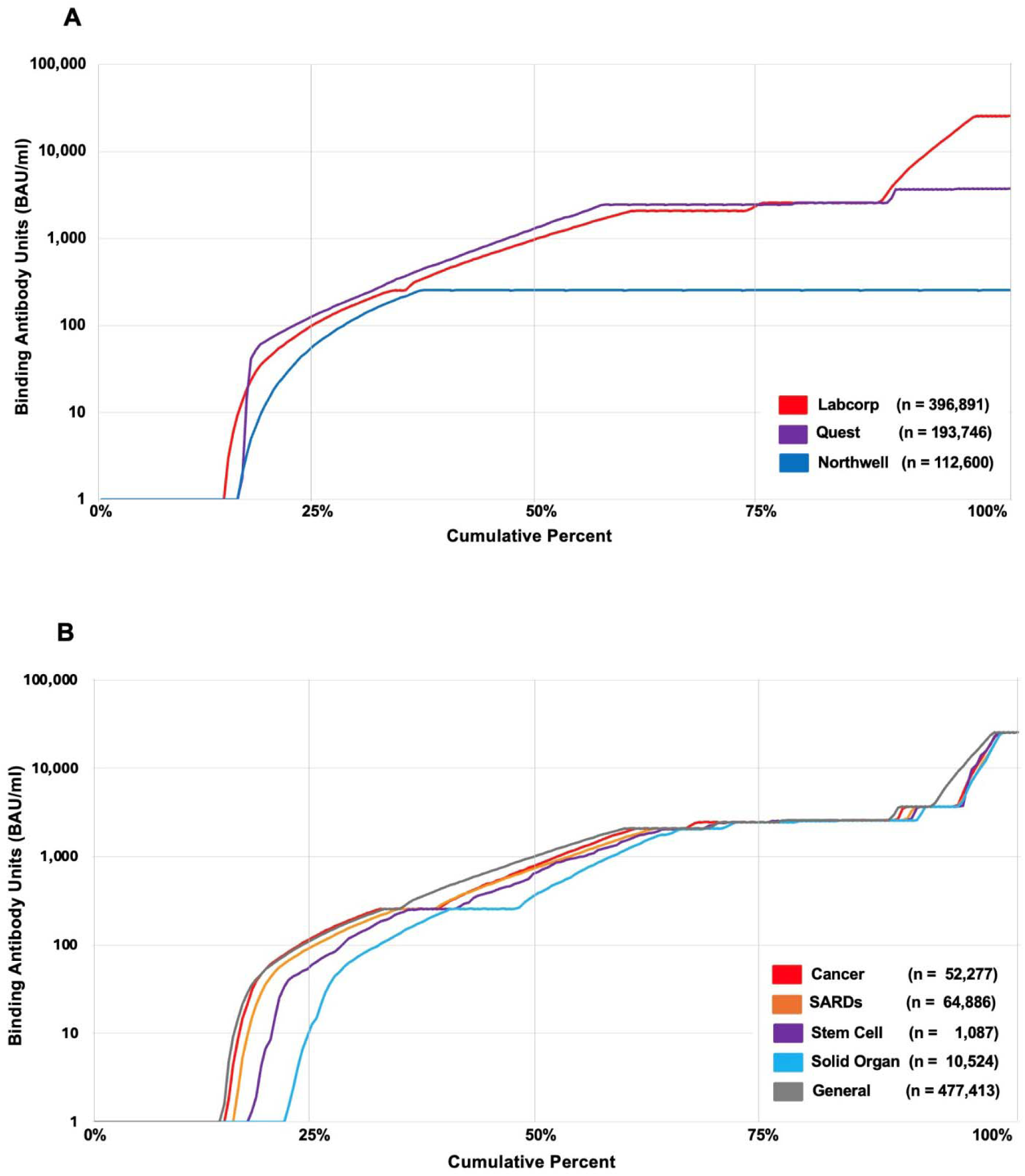
Cumulative semilog ordinal plots of unique testing events for semi-quantitative SARS-CoV-2 Anti-S serology by source laboratory and immunocompromised condition. The x-axis shows the cumulative percentage of testing events. The y-axis shows the harmonized binding arbitrary units (BAU), per the logic given in Table 1. The legends give the total testing events for each curve. (A) Cumulative ordinal plots by source laboratory. (B) Cumulative ordinal plots by indexed immunocompromised condition; a general population of non-immunocompromised persons is shown for comparison.

We next examined semi-quantitative serologic anti-S test data based on indexed immunocompromised status (Figure 2B). Most serologic tests were performed on a comparison general population: 13% exhibited harmonized BAU test results below the reference threshold for negative; from 13% to 52% the test results were in a submaximal range from 1-2500 BAU; and above 52%, test results were maximal at ≥2500 BAU. For the 4 immunocompromised cohorts, 14% of specimens from patients with cancer had negative test results, compared to 15% for patients with SARDs, 17% for patients who had undergone stem cell transplant, and 21% for patients who had undergone solid organ transplant patients. The cumulative distributions in the submaximal dynamic range lagged similarly for the 2 transplant cohorts. Once test results plateaued in the ≥2500 BAU range, the curves were largely similar, although there was separation of the general population cohort in the rise from 2500 to ≥25 000 BAU.

## Discussion

Harmonization of laboratory testing is a foundational principle for the practice of laboratory medicine (29). However, while analytical standardization of results from clinical laboratory in vitro diagnostics measurement procedures can be achieved through metrological traceability of results to certified reference materials or reference measurement procedures, (30), specifics regarding the naming of tests for ordering and reporting is the prerogative of individual laboratories. In this instance, laboratories were implementing testing in response to a pandemic, with new tests and platforms being approved at a rapid rate, for an unknown level of demand for testing capacity. The fact that 4 source laboratories generated 87 different test names and 41 different ways to report “positive” or “negative” provides an important learning opportunity relevant to preparedness for future pandemics, when testing platforms and regulatory approvals thereof may be changing rapidly. Beyond that, we consider that these strategies are of general relevance to harmonization of real-world laboratory data.

While the LOINC coding system aims to standardize both the test name and the specifics of the assay platform (31), the above pluralities show that there is insufficient structured content in LOINC to permit unsupervised harmonization of test names and even qualitative test results (32). Semi-quantitative serological testing is a particular challenge, since until a serological assay is harmonized to an international standard by the manufacturer, results are given in BAU, specific to each manufacturer. For vaccine-preventable infectious disease pathogens such as measles, hepatitis B virus, mumps, rubella and varicella, the World Health Organization (WHO) standard is in place for reporting of test results (33–35). Such standardization was not generally achieved for commercial SARS-CoV-2 serological testing, despite publications for doing so from both industry and the SeroNet consortium (36,37).

Moreover, despite extensive publications reporting on the serological responses of patient populations to natural SARS-CoV-2 infection and COVID-19 vaccination, US guidelines remain silent on the interpretation of semi-quantitative serologic testing. The August 29, 2024 recommendations of the Centers for Diseases Prevention and Control (CDC, 28) are essentially unchanged from the 2020 recommendations: SARS-CoV-2 serologic testing does not diagnose current infection and is primarily used for public health surveillance and epidemiologic purposes. Per the CDC recommendations, the presence of SARS-CoV-2 antibodies may aid in fulfilling the case definition for multisystem inflammatory syndrome in children (MIS-C) and adults (MIS-A) and may help in distinguishing an immune response from natural infection versus vaccination.

There remains no further CDC comment the clinical utility of such testing, such as assessing need for vaccination of patients with immunocompromising conditions, although such testing may be of benefit (27). Barriers to establishing such utility include the differing assay cut-offs for negative and positive results; the impact of SARS-CoV-2 variants on host immune response; the absence of established correlates of protection, and lack of clarity on the role of T cells in preventing severe disease (25).

The decline in the ordering of SARS-CoV-2 serologic testing for calendar years 2023 and 2024 documented in this report underscores the need for using existing datasets to gain insights into the potential clinical utility of such testing, especially in vulnerable populations. The CRWDi data resource constitutes a large harmonized dataset for study of patients with immunocompromising conditions. The methodology described in this report, and the infrastructure that has been built for harmonization of all SARS-CoV-2 laboratory tests, make such research possible.

Several observations can be made from the data presented herein. First, the increase in qualitative serologic testing in May, June and July 2020 is likely a reflection of the serosurveillance studies performed in the early months of the pandemic (38), including healthcare workforce studies (28). The substantial decrease in calendar years 2023 and 2024 of both laboratory-based NAAT and serology testing likely reflects utilization of over-the-counter antigen testing for home-based COVID-19 diagnosis, and the lack of indication for testing SARS-CoV-2 antibody levels in clinical practice. This is despite COVID-19 accounting for 1.6% (49,303) of the 3 million total deaths in the US in 2023 and remaining the tenth most common cause of death, particularly in the elderly and in vulnerable populations (39).

Second, the preponderance of COVID-19 illness-related ICD-10 codes being provided by administrative claims data underscores the value of harmonizing SARS-CoV-2 laboratory testing with administrative data to understand test utilization. Third, while the performance or not of autodiliution of serologic specimens by the source laboratories may have differed (Figure 2A), the overall similarities of the semilog ordinal curves support the premise that the source laboratories are testing populations of similar composition. This provides justification for merging of the respective datasets for further studies. Fourth, Figure 2B documents that specimens from patients with stem cell transplantation and especially solid organ transplantation exhibit both a higher percentage of negative results, and a lag in the number of specimens that reach maximal values over the dynamic range of the semi-quantitative assay, when compared with the general population and with specimens from patients with cancer or SARDs. It will be for future studies to determine the characteristics of patients from whom these specimens were obtained, and especially whether negative or submaximal semiquantitative test results can serve as correlates of protection regarding outcomes.

Limitations of this study are as follows. First, this is a methodological report of harmonization of unique test results, and is not an actual study of the clinical utility of the testing. This includes the fact that vaccination status or history of SARS-CoV-2 infection were not part of our analysis. Second, variable autodilution practices for anti-S serological results (Figure 2A) may limit interpretation of maximal test results. However, we consider that patients with negative or submaximal antibody levels are of greater interest, on the basis of an absent or lower humoral response. Third, the indexing of patients by condition (Figure 2B) does not consider the impact of immunomodulating therapy that these patients may have received. It will be of importance to include the immunosuppressive regimes being received by patients as a covariable in future studies using these data.

## Conclusion

This report provides a generalizable methodology for harmonizing qualitative and semiquantitative laboratory testing data for a vaccine-preventable pathogen, in this case SARS-CoV-2. Both diagnostic and serological testing data from multiple laboratory sources can be successfully harmonized with the broad geography of real-world administrative claims and vital status data. For CRWDi, this also includes harmonization with cancer diagnostic, therapeutic, and longitudinal outcomes data from the SEER registry system, representing a particular enhancement of this publicly available resource. This report also demonstrates the specific need for harmonization of real-world laboratory testing data when performed during the rapidly evolving time frame of a pandemic.

## Abbreviations

CDC: Centers for Disease Control and Prevention
CPT: Current Procedural Terminology
CRWDi: COVID-19 Real World Data infrastructure
Ct: cycle threshold
HCPCS: Healthcare Common Procedure Coding System
HSC: Hematopoietic stem cell
HVID: HealthVerity de-identified patient identification number
HVM: HealthVerity Marketplace
ICD-10: International Classification of Disease, 10^th^ Revision
LOINC: Logical Observation Identifier Names and Codes
NAAT: Nucleic Acid Amplification Test
NCI: National Cancer Institute
NDC: National Drug Code
PHI: Personalized health information
PPRL: Privacy preserving record linkages
SARDs: systemic autoimmune and rheumatic diseases
SEER: Surveillance, Epidemiology, and End Results program
US: United States
WHO: World Health Organization

## References

1. Litt D, Slack MPE, Nakamura T, et al. Evaluation of the World Health Organization Global Invasive Bacterial Vaccine-Preventable Disease (IB-VPD) Surveillance Network’s Laboratory External Quality Assessment Programme, 2014-2019. J Med Microbiol. 2023; 72(1): 001644. DOI: 10.1099/jmm.0.001644.

2. Marcenac P, McCarron M, Davis W, et al. Leveraging International Surveillance Systems and Programs during the COVID-19 pandemic. Emerging Infect Dis. 2022; 28: S26–S33. DOI: 10.3201/eid2813.212248.

3. Haendel MA, Chute CG, Bennett TD, et al. The National COVID Cohort Collaborative (N3C): Rationale, design, infrastructure, and deployment. J Am Med Inform Assoc. 2021; 28: 427–443. DOI: 10.1093/jamia/ocaa196.

4. Liew JW, Bhana S, Costello W, et al. The COVID-19 Global Rheumatology Alliance: evaluating the rapid design and implementation of an international registry against best practice. Rheumatology 2021; 60: 353–358.

5. Scobie HM, Johnson AG, Suthar AB, et al. Monitoring incidence of COVID-19 cases, hospitalizations, and deaths, by vaccination status – 13 U.S. jurisdictions, April 4 – July 17, 2021. Morbidity Mortality Weekly Report, September 17, 2021; 70(37): 1284–1290.

6. Song S, Madewell ZJ, Liu M, et al. A systematic review and meta-analysis on the effectiveness of bivalent mRNA booster vaccines against Omicron variants. Vaccine. 2024; 42: 3389–3396.

7. Bonanni P, Ceddia F, Dawson R. A call to action: Current challenges and considerations for COVID-19 vaccination in immunocompromised populations. J Infect Dis, 2023:228 (Suppl 1): S70–S76. DOI: 10.1093/infdis/jiad150.

8. Antinori A, Bausch-Jurken M. The burden of COVID-19 in the immunocompromised patient: implications for vaccination and needs for the future. J Infect Dis 2023; 228: S4–S12.

9. Bonanni P, Canton R, Gill D, et al. The role of serology testing to strengthen vaccination initiatives and policies for COVID-19 in Europe. COVID, 2021; 1: 20–38. DOI: 10.3390/covid1010004.

10. Hayden MK, El Mikati IK, Hanson KE, et al. Infectious Diseases Society of America guidelines on the diagnosis of COVID-19: Serologic testing. Clin Infect Dis 2024. DOI: 10.1093/cid/ciae121.

11. Pfaff ER, Girvin AT, Crosskey M, et al. De-black-boxing health AI: demonstrating reproducible machine learning compatible phenotypes using the N3C-RECOVER Long COVID model in the *All of Us* data repository. J Am Med Informatics Assoc. 2023; 30: 1305–1312.

12. Zang C, Hou Y, Schenck EJ, et al. Identification of risk factors of Long COVID and predictive modeling in the RECOVER EHR cohorts. Commun Med (Lond). 2024 Jul 11; 4(1): 130. DOI: 10.1038/s43856-024-00549-0.

13. Vinson AJ, Schissel M, Anzalone AJ, et al. The prevalence of post-acute sequelae of COVID-19 in solid organ transplant recipients: Evaluation of risk in the National COVID Cohort Collaborative (N3C). Am J Transpl 2024; S1600-6135(24)00370-8. DOI: 10.1016/j.ajt.2024.06.00114.

14. Preiss A, Bhatia A, Aragon LV, et al. Effect of Paxlovid treatment during acute COVID-19 on Long COVID onset: An EHR-based target trial emulation from the N3C and RECOVER consortia. PLoS Med 2025; 22(9): e1004711. DOI: 10.1371/journal.pmed.1004711.

15. Anderson E, Powell M, Yang E, Kar A, Leung TM, Sison C, et al. Factors associated with immune responses to SARS-CoV-2 vaccination in individuals with autoimmune diseases. JCI Insight 2024; 9(13): e180750. DOI: 10.1172/jci.insight.180750.

16. Skarbinski J, Elkin EP, Ziemba YC, Kazemian E, Wilson BM, Siddiqui H, Schleicher CB, Hsiao CA, Nugent JR, Reckamp KL, Merchant A, Crawford JM, Zidar DA, Kushi LH, Figueiredo JC. (2025) COVID-19 vaccine uptake and effectiveness among persons with cancer on chemotherapy or immunotherapy during two time periods in four health systems in the United States. JAMA Oncology 2025 Sep 21; 11(9): 999–1010 (ePub: Jul 17:e252020). DOI: 10.1001/jamaoncol.2025.2020.

17. Ziemba YC, Elkin EP, Kazemian E, Wilson BM, Siddiqui H, Schleicher CB, Hsiao CA, Zidar DA, Kushi LH, Figueiredo JC, Skarbinski J, Crawford JM. (2025) COVID-19 booster vaccine uptake and effectiveness among people with systemic autoimmune rheumatic diseases. J Rheumatology, 2025 Aug 1:jrheum.2025-0535. DOI: 10.3899/jrheum.2025-0535.

18. Centers for Disease Prevention and Control. Overview of Testing for SARS-CoV-2. https://www.cdc.gov/covid/hcp/clinical-careoverview-testing-sars-cov-2.html. Accessed December 14, 2025.

19. Titova E, Kan VW, Lozy T, et al. Humoral and cellular immune responses against SARS-CoV-2 post-vaccination in immunocompetent and immunocompromised cancer populations. Microbiol Spectrum. 2024, 12(3):e0205023. DOI: 10.1128/spectrum.02050-23.

20. Haselbeck AH, Im J, Prifti K, et al. Serology as a tool to assess infectious disease landscapes and guide public health policy. Pathogens. 2022, 11: 732. DOI: 10.3390/pathogens11070732.

21. Gertosio C, Licari A, De Silvestri A, et al. Efficacy, immunogenicity, and safety of available vaccines in children on biologics: A systematic review and meta-analysis. Vaccine. 2022; 40: 2679–2695.

22. Rondaan C, Furer V, Heijstek MW, et al. Efficacy, immunogenicity and safety of vaccination in adult patients with autoimmune inflammatory rheumatic diseases: a systematic literature review for the 2019 update of EULAR recommendations. RMD Open 2019; 5: e001030. DOI: 10.1136/rmdopen-2019-001035.

23. Rubin LG, Levin MJ, Ljungman P, et al. 2013 IDSA clinical practice guideline for vaccination of the immunocompromised host. Clin Infect Dis 2014; 58: 309–318.

24. Hostetler HP, Neely ML, Lydon E, et al. Immunity to varicella, measles, and mumps in patients evaluated for lung transplantation. Am J Transplant 2021; 21: 2864–2870.

25. Patel C, Wang X, Giles M, et al. The role of serological testing for vaccination with a focus on immunocompromised patients: a synthesis of current evidence. Vaccine, 2025; 63: 127659. DOI: 10.1016/vaccine.2025.127659.

26. Crawford JM, Penberthy L, Pinto LA, et al. COVID-19 Real World Data Infrastructure: A big data resource for study of the impact of COVID-19 in patient populations with immunocompromising conditions. Open Forum Infectious Diseases. 2025 Jan 23; 12(1):ofaf021. DOI: 10.1093/ofid/ofaf021.

27. Kaufman HW, Letovsky S, Meyer WA III, et al. SARS-CoV-2 spike-protein targeted serology test results and their association with subsequent COVID-19-related outcomes. Front. Public Health 2023; 11:1193246. DOI: 10.3389/fpubh.2023.1193246.

28. Moscola J, Sembajwe G, Jarrett M, Farber B, Chang T, McGinn T, Davidson KW, and the Northwell Health COVID-19 Research Consortium (Becker LB, Breining DA, Moline J, Butler MJ, Cookingham J, Dominello AJ, Falzon L, Foroughi C, Johnson JC, Mogavero JN, Monane R, Vicari F, Crawford JM, Fox SS, Juretschko S, Schleicher CB, Hutchins CJ, McPhillips KM, Saraya A). (2020) Prevalence of SARS-CoV-2 antibodies in health care personnel in the New York City area. J Am Med Assoc, 6 Aug 2020, Research Letter published on-line. DOI: 10.1001/jama.2020.14765.

29. Plebani M. Harmonization in laboratory medicine: more than clinical chemistry? Clin Chem Lab Med 2018; 56(10: 1579–1586.

30. Miller WG, Greenberg N. Harmonization and standardization: Where are we now? J Appl Lab Med, 2021 March 1; 6(2): 510–521. DOI: 10.1093/jalm/ifaa189.

31. LOINC® from Regenstrief. Home. LOINC®. 2021. http://www.LOINC.org/. Accessed December 15, 2025.

32. Cholan RA, Pappas G, Rehwoldt G, et al. Encoding laboratory testing data: case studies of the national implementation of HHS requirements and related standards in five laboratories. J Am Med Informatics Assoc, 2022; 29: 1372–1380.

33. Dimech W, Panagiotopoulos L, Francis B, et al. Evaluation of eight anti-rubella virus immunoglobulin g immunoassays that report results in international units per milliliter. J Clin Microbiol, 2008; 46: 1955–1960.

34. Hesketh I, Charlett A, Farrington P, et al. An evaluation of nine commercial EIA kits for the detection of measles specific IgG. J Virol Methods 1997; 66: 51–59.

35. Huzly D, Hanselmann I, Neumann-Haefelin D, Panning M. Performance of 14 rubella immunoassays on samples with low positie or negative haemagglutination inhibition results. J Clin Virol 2016; 74: 13–18.

36. Freeman J, Conklin J. Standardization of two SARS-CoV-2 serology assays to the WHO 20/136 human standard reference material. J Virol Meth 2022; 300: 114430. DOI: 10.1016/j.jviromet.2021.114430.

37. Kemp TJ, Quesinberry JT, Cherry J, et al. Selection, characterization, calibration, and distribution of the U.S. Serology Standard for Anti-SARS-CoV-2 antibody detection. J Clin Microbiol 2022; 60:00995-22. DOI: 10.1128/jcm.00995-22.

38. Angulo FJ, Finelli L, Swerdlow DL. Estimation of US SARS-CoV-2 infections, symptomatic infections, hospitalizations, and deaths using seroprevalence surveys. JAMA Network Open. 2021;4(1):e2033706. DOI: 10.1001/jamanetworkopen.2020.33706.

39. Ahmad FB, Cisewski JA, Anderson RN. Leading causes of death in the US, 2019-2023. JAMA. 2024; 332: 957–958.

