## Supplemental Material for "Harmonization of Real-World Data for Vaccine-Preventable Infectious Diseases: Integration of SARS-CoV-2 Diagnostic and Serologic Data from Multiple Sources"

#### Supplemental Table S1. SARS-CoV-2 test name recoding to test category

| NAAT (n=27 test names) | Serology_Qualitative (n=34 test names) | Serology_Semi-Quantitative (n=26 test names) |
|---|---|---|
| SARS-CoV-2 RNA, QUALITATIVE REAL-TIME RT-PCR | SARS-CoV-2 Antibody, IgG | SARS CoV 2 IGM |
| Pan-SARS RNA: | SARS COV 2 IGM | SARS COV2,IGG,SPKE,SEMIQN |
| Overall Result: | Abbott SARS-CoV-2 Ab, IgG | SARS COV2 AB,TOT SPIKE SEMI QN |
| SARS-CoV-2, NAA | SARS-CoV-2 Antibody, IgG | SARS CoV2,IgG,Spke,SemiQN |
| SARS-Cov2 | ANTI-SARS-COV-2 ABS | COVID-19 Total IgG Measurement |
| SARS-CoV-2 | SARS COV 2 AB (IGG) | SARS COV 2 AB, TOTAL SPIKE SEMI QN |
| COVID-19 PCR | SARS CoV 2 AB (IgG) | COVID-19 Nucleocapsid Antibody Measurement |
| COVID-19 NAA, Saliva | SARS COV 2 AB (IGM) | SARS CoV 2 AB IGG |
| SARS-CoV-2 RNA, Qualitative Re | SARS COV 2 AB IGG | ANTI-SARS-COV-2-S (SPIKE) |
| SARS COV2 RNA | COVID-19 Nucleocapsid Antibody Interpretation | SPIKE ANTI SARS COV2 ABS |
| SARS COV-2 RNA: | DiaSorin SARS-CoV-2 Ab, IgG | SARS-CoV-2 Semi-Quant Spike Ab |
| SARS-CoV RNA, Qualitative Real-Time RT-PCR | Euroimmun SARS-CoV-2 Ab, IgG | SARS COV2,TOT,SPKE,SEMIQN |
| PAN-SARS RNA: | SARS CoV 2 (COVID 19) AG, POC, FIA (QUIDL) | SARS-CoV-2 Semi-Quant Total Ab |
| SARS CoV 2 RNA: | COVID-19 Spike Antibody Measurement | COVID-19 Spike Antibody Measurement |
| OVERALL RESULT: | COVID-19 Total IgG Interpretation | SARS-CoV-2 Semi-Quant IgG Ab |
| CRSP SARS COV 2 REAL TIME (RT) PCR DIAG ASSAY | SARS-CoV-2 IgG | SARS COV2 AB (IGG) SPIKE), SEMI QN |
| SARS-CoV-2 PCR | SARS CoV 2 IGM | SARS CoV2,IgG,Spike,SemiQN |
| SARS COV 2 RNA, RT PCR | SARS-CoV-2 Ab, Nucleocapsid | SARS-CoV-2 Spike Ab Dilution |
| SARS CoV 2 REAL TIME (RT) PCR, DIAGNOSTIC | SARS CoV 2 Ab (IgM) | SARS CoV2,Tot,Spke,SemiQN |
| SARS-CoV-2 RNA | SARS CoV 2 Ag, POC NUCLEOCAPSID,QL,(ABNX) | SARS CoV 2 Ab (IgG) |
| SARS CoV 2 RNA | COVID-19 Spike Antibody Interpretation | SARS COV 2 AB (IGG) |
| SARS-CoV-2 RNA: | SARS CoV 2 Ab (IgG) | SARS-CoV-2 Ab, Nucleocapsid |
| SARS CoV 2 RNA, RT PCR | SARS-CoV-2 IgG, Dried Blood | SARS COV 2 AB IgG SPIKE SEMI QN |
| SARS COV 2, PCR | SARS CoV 2 AB (IgM) | SARS-CoV-2 Spike Ab Interp |
| Pan-SARS RNA | SARS COV 2 AB IGG | SARS COV 2 IGM |
| SARS CoV RNA | SARS-CoV-2 Spike Ab Interp | SARS COV 2 SEROLOGY COVID 19 AB IGG |
| SARS CoV2 RNA | SARS-CoV-2 Antibody, IgA | |
| | SARS-CoV-2 Antibodies | |
| | SARS COV 2 AB (IGG) SPIKE, SEMI QN | |
| | 39504 SARS COV 2 IGG | |
| | COVID-19 Total IgG Measurement | |
| | COVID 19 IgG | |
| | SARS CoV 2 Ab IgG | |
| | SARS CoV 2 ANTIBODY IgG | |

Test names were as contributed by 4 source laboratories. Recoding of NAAT was self-evident. For Serology, the correct categorization and sub-categorization of each test name was verified with the 2 contributing commercial laboratories (Labcorp, Quest) and with Northwell. The serology sub-categories were antigen tested (total, S, N) and antibody class.

### Supplemental Table S2. SARS-CoV-2 recoding of test results

| Result Submitted by Laboratory | Data vendor | Recoded result | Count |
|---|---|---|---|
| LDTNOT | Labcorp | Negative | 133 601 |
| N | Labcorp | Negative | 412 605 |
| Not Detected | Labcorp | Negative | 1 480 290 |
| Negative | Labcorp | Negative | 724 754 |
| N | Quest Diagnostics | Negative | 14 |
| Not detected | Quest Diagnostics | Negative | 1 |
| NOTDETECTED | Quest Diagnostics | Negative | 1 |
| Not Detected | Quest Diagnostics | Negative | 266 996 |
| NOT DETECTED | Quest Diagnostics | Negative | 1 224 167 |
| Non-Reactive | Quest Diagnostics | Negative | 225 |
| <1.00 NEGATIVE | Quest Diagnostics | Negative | 1 |
| <1.00 Negative | Quest Diagnostics | Negative | 1 |
| Negative^ | Quest Diagnostics | Negative | 486 |
| Negative | Quest Diagnostics | Negative | 157 601 |
| NEGATIVE | Quest Diagnostics | Negative | 703 207 |
| NEG | Quest Diagnostics | Negative | 4 |
| NEGATUVE | Quest Diagnostics | Negative | 1 |
| nNegative | Quest Diagnostics | Negative | 1 |
| not Detected | Private Source 74 | Negative | 4 |
| NOT DETECTED | Private Source 74 | Negative | 10 029 |
| Not Detected | Private Source 74 | Negative | 693 227 |
| SARS-CoV-2 Not Detected | Private Source 74 | Negative | 150 109 |
| NEGATIVE | Private Source 74 | Negative | 38 |
| Negative | Private Source 74 | Negative | 2 326 |
| Covid-19 Virus NOT detected | Private Source 74 | Negative | 12 |
| NEGATIVE <1.00 | Northwell | Negative | 1 |
| NEGATIVE | Northwell | Negative | 452 855 |
| NOT DETECTED | Northwell | Negative | 13 |
| NEG | Northwell | Negative | 3 |
| LDTDET | Labcorp | Positive | 13 833 |
| P= | Labcorp | Positive | 32 137 |
| P | Labcorp | Positive | 40 652 |
| POS | Labcorp | Positive | 150 |
| Positive | Labcorp | Positive | 598 570 |
| Detected | Labcorp | Positive | 159 070 |
| P | Quest Diagnostics | Positive | 4 |
| Positive^ | Quest Diagnostics | Positive | 170 |
| Positive | Quest Diagnostics | Positive | 53 941 |
| POSITIVE | Quest Diagnostics | Positive | 168 889 |

| | | | |
|---|---|---|---|
| Detected | Quest Diagnostics | Positive | 27 913 |
| DETECTED | Quest Diagnostics | Positive | 153 181 |
| POS | Quest Diagnostics | Positive | 4 |
| Reactive | Quest Diagnostics | Positive | 66 |
| DTECTED | Quest Diagnostics | Positive | 1 |
| > or = 1.00 Positive | Quest Diagnostics | Positive | 1 |
| POSITIVE | Private Source 74 | Positive | 2 |
| Positive | Private Source 74 | Positive | 464 |
| Positive SARS-CoV-2 | Private Source 74 | Positive | 12 763 |
| Detected | Private Source 74 | Positive | 9 |
| DETECTED | Private Source 74 | Positive | 1 266 |
| Positive for 2019-nCoV | Private Source 74 | Positive | 71 761 |
| Presumptive Positive for 2019-nCoV | Private Source 74 | Positive | 2 200 |
| Presumptive Pos. for 2019-nCoV | Private Source 74 | Positive | 123 |
| Presumptive Positive 2019-nCoV | Private Source 74 | Positive | 3 |
| Covid-19 Virus detected | Private Source 74 | Positive | 3 |
| POSITIVE | Northwell | Positive | 207 872 |
| POSTIIVE | Northwell | Positive | 1 |
| POS | Northwell | Positive | 2 |
| DETECTED | Northwell | Positive | 5 |
| LT0.8 | Labcorp | <0.8 | 2 497 |
| LT13 | Labcorp | <13.0 | 84 |
| SAR250 | Labcorp | >250.0 | 6 |
| TROPHI | Labcorp | >250.00 | 106 |
| SARSQE | Labcorp | >2500.0 | 83 |
| GT25TH | Labcorp | >25000 | 15 404 |
| GT800 | Labcorp | >800.0 | 650 |

Test results were as contributed by all source laboratories. Negative and positive result texts from the 2 named commercial laboratories (Labcorp, Quest) and Northwell were verified with the source laboratories. Negative and positive results from the unnamed commercial laboratory ("private") were considered self-evident. The lowest frequency sequences (less than 10 results, representing 22 text sequences) included spelling errors. But even with a threshold of 1000 or more results with a specific alphanumeric sequence, there were still 37 different text sequences to communicate "negative" or "positive" test outcomes. A reference recode table was created to standardize resulting of a negative or positive qualitative test result. Separately, one source laboratory (Labcorp) reported a subset of alphanumeric qualitative results with imputed numerical values. In the recoding step, we chose to retain this imputed numerical information for the semi-quantitative serologic dataset, as shown in the last 7 rows of Supplemental Table 3. We found that these alphanumerical results accompanied replicate "negative" or "positive" qualitative results for the same testing event, so that a true qualitative result also existed for this limited subset of test results.

**Supplemental Table S3. Manufacturer Assay Platforms for SARS-CoV-2 Serologic Testing**

| **Manufacturer** | **Platform** | **Methodology** |
|---|---|---|
| Abbott Diagnostics (Chicago, IL, USA) | Architect | CMIA |
| Diasorin (Stillwater, MN, USA) | Liaison | CLIA |
| Euroimmun Medizinsche Labordiagnostika (Lubeck, Germany) | Inova Quantalyser | ELISA |
| Ortho Diagnostics (Raritan, NJ, USA) | Vitros | CLIA |
| Roche Diagnostics (Indianapolis, IN, USA) | Elecsys Cobas | ECLIA |
| Siemens Healthineers (Tarrytown, NY, USA) | Atellica | CLIA |

Abbreviations: CLIA, chemiluminescent immunoassay; CMIA chemiluminescent microparticle immuno-assay; ECLIA, electrochemiluminescence immunoassay; ELISA, enzyme-linked immunosorbent assay.

### Supplemental Table S4: Relevant ICD-10 codes accompanying the SARS-CoV-2 Test Order

| | NAAT | Serology Qualitative | Serology Semi-quantitative | Total |
|---|---|---|---|---|
| **Asymptomatic screen** | **157 359** | **342 065** | **85 477** | **584,901** |
| Z00.00 Encounter for general adult medical exam w/o abnormal findings | 3 858 | 114 938 | 27 636 | 146 432 |
| Z00.01 Encounter for general adult medical exam w abnormal findings | 1 214 | 39 499 | 12 467 | 53 180 |
| Z00.129 Encounter for routine child health exam w/o abnormal findings | 310 | 13 537 | 2 353 | 16 200 |
| Z01.812 Encounter for preprocedural laboratory examination | 6 663 | 1 965 | 271 | 8 899 |
| Z01.818 Encounter for other preprocedural examination | 18 855 | 5 606 | 1 762 | 26 223 |
| Z01.84 Encounter for antibody response examination | 6 850 | 46 741 | 15 071 | 68 662 |
| Z11.52 Encounter for screening for COVID-19 | 25 809 | 10 775 | 10 802 | 47 386 |
| Z11.59 Encounter for screening for other viral diseases | 92 610 | 92 022 | 10 951 | 195 583 |
| Z13.0 Encounter screen for diseases of the blood/blood-forming organs/immune system | 244 | 9 217 | 2 691 | 12 152 |
| Z13.9 Encounter for screening, unspecified | 946 | 7 765 | 1 473 | 10 184 |
| **Contact** | **1 127 777** | **451 738** | **44 849** | **1 624 364** |
| Z03.818 Encounter for observation for suspected exposure to other biologic agents ruled out | 79 263 | 66 323 | 6 885 | 152 471 |
| Z20.822 Contact with and (suspected) exposure to COVID-19 | 429 456 | 39 307 | 14 945 | 483 708 |
| Z20.828 Contact with and exposure to other viral communicable diseases | 607 690 | 336 040 | 22 204 | 965 934 |
| Z20.89 Contact with and exposure to other communicable diseases | 11 368 | 10 068 | 815 | 22 251 |
| **Symptoms or disease condition** | **211 791** | **234 237** | **54 426** | **500 454** |
| B34.2 Coronavirus infection, unspecified | 5 103 | 7 453 | 1 449 | 14 005 |
| B34.9 Viral infection, unspecified | 20 401 | 14 691 | 1 457 | 36 549 |
| J02.9 Acute pharyngitis, unspecified | 18 032 | 3 375 | 375 | 21 782 |
| J06.9 Acute upper respiratory infection, unspecified | 25 537 | 6 485 | 775 | 32 797 |
| J30.9 Allergic rhinitis, unspecified | 1 157 | 6 044 | 2 041 | 9 242 |
| R05 Cough | 38 105 | 21 291 | 1 080 | 60 476 |
| R05.9 Cough, unspecified | 12 224 | 2 148 | 1 249 | 15 621 |
| R06.02 Shortness of breath | 8 167 | 7 166 | 1 418 | 16 751 |
| R50.9 Fever, unspecified | 25 113 | 8 945 | 1 321 | 35 379 |
| R53.81 Other malaise | 1 542 | 8 133 | 2 290 | 11 965 |
| R53.82 Chronic fatigue, unspecified | 424 | 6 985 | 2 953 | 10 362 |
| R53.83 Other fatigue | 9 463 | 54 380 | 15 049 | 78 892 |
| R68.89 Other general symptoms and signs | 7 654 | 11 142 | 2 766 | 21 562 |
| U07.1 COVID-19 | 37 783 | 62 554 | 13 858 | 114 195 |
| Z86.16 Personal history of COVID-19 | 1 086 | 13 445 | 6 345 | 20 876 |
| **Grand Total** | **1 496 927** | **1 028 040** | **184 752** | **2 709 719** |

COVID-19 relevant ICD-10 codes accompanying laboratory test orders are shown, as available in CRWDi. As these are based on transmission of this data field to HealthVerity for harmonization into CRWDi, there may be missing data and the numbers may not reflect all the ICD-10 codes present in the source laboratory information systems. Relevance was defined as ICD-10 codes pertaining to screening (including encounter), contact, respiratory illness, and/or symptomatology that might arise from COVID-19 illness.
